# Preference and Perception of Indian Caregivers Towards Formal Long–Term Supportive Services – A Mixed–Methods Study

**DOI:** 10.1101/2023.12.01.23299284

**Authors:** Subharati Ghosh, Arvind Mathur

## Abstract

**Purpose of the Study:** In India, the rate of population aging, the greater burden of chronic disease-related care, and smaller families question the sustainability of traditional family based, home long term support and services (LTSS). However, little is still known about Indians’ perception of formal LTSS.

**Design and Methods:** A mixed-methods design of family caregivers for older adults from Jodhpur, Rajasthan (n=30 in-depth interviews; n=100 quantitative survey). Inductive qualitative data analysis identified emergent themes about perceptions of either informal or formal LTSS. Caregivers self-reported which common LTSS they needed in a quantitative survey. Multivariable Poisson and logistic regression models were used to estimate the average total number of LTSS and probabilities of self reporting wanting individual LTSS, respectively.

**Results:** The central theme was a negative perception of formal LTSS, especially the idea of paid helpers. A second theme served as the rationale for the first theme: caregivers reported a “duty” to provide care to one’s family that “others” and those “doing it for money” could not meet. Caregivers reported on average 2.8 LTSS needs of 10 options. Formal LTSS, like home-health care assistance with instrumental activities of daily living, were least frequently reported; caregiver education and self-care activities were the most reported.

**Implications:** Despite providing intensive amounts of informal LTSS and care for their family members, Indian caregivers consistently reported disinterest in using formal LTSS alternatives in qualitative and quantitative data. Caregivers reported a stronger desire for services that support their ability to carry out their caregiving roles.

## Introduction

In lower- and middle-income countries, the care needs of older persons nearly always fall to family members alone, overwhelmingly to women.(Apt & Gricco, 1994; Wachter, 1997) The number of older persons to care for is falling on fewer and fewer shoulders: globally, there will be 24 persons aged 65+ for everyone aged 15-64 in 2050 compared to 11:1 in 2010. (“World Population Prospects: The 2015 Revision,” 2015) Although low and middle-income countries are home to over 2/3rds of the world’s population aged 65+, it remains unclear what are the best approaches – either formal health care or informal family care systems – to support this unprecedentedly large number of older adults in these settings. In particular, there are crucial gaps in our understanding of individuals’ perceptions of their caregiving roles and their preferences for supportive interventions to facilitate carrying out their ideal perception of care for older adult family members.

Older adults in India often lack access to health care, especially to geriatricians, which remains a rare specialty throughout the country.(Lloyd-Sherlock, 2000, 2002; Shrestha, 2000) Generally speaking, despite efforts to improve access to health care and for the government-sponsored health care system to address the health of Indians (Vikram Patel et al., 2015), much of the health care Indian adults receive is from private providers of varying quality,(Powell-Jackson, Acharya, & Mills, 2013) and with significant out-of-pocket expenditures. (Balarajan, Selvaraj, & Subramanian, 2011; Duran, Kutzin, & Menabde, 2014; Kumar et al., 2011; Prinja, Aggarwal, Kumar, & Kanavos, 2012; Shankar Prinja et al., 2012; Reddy et al., 2011; van Doorslaer et al., 2006) Non-communicable diseases are associated with significant health care utilization and out-of-pocket expenditures. (Pati et al., 2014)

Formal care infrastructure, such as long-term or residential care facilities (Lloyd-Sherlock, 2002) are often scarce. In India, historically, the family is the most socially acceptable place to age (Rajan & Kumar, 2003; Jamuna, 2003). However, there is some indication that these patterns are steadily changing, as evidenced by a steady increase in nuclear living arrangements among older adults, and the number of older adults living alone or only with their spouse has increased over time. These patterns naturally raise the question, “Where are older adults going to age and who is going to care for the elderly in India?” Despite a perceived cultural stigma associated with formal long-term care use, this evidence remains more anecdotal than empirical. Many older adults living in a formal, residential “old age home” reported having been threatened by their children with being kicked out of their home and taken to such a home.(Kalavar, Jamuna, & Ejaz, 2013)

The broader study drew largely from Pearlin’s stress-coping model(Leonard I. Pearlin, Mullan, Semple, & Skaff, 1990), which argues that stressors, of which caregiving is one, can affect physical health and psychological well-being. However, the relationship between stress and caregiving is moderated by caregiving support. A recent study on long-term care of adults with Autism in India used a similar theoretical model to identify factors associated with long-term planning (Ghosh, 2022). In the absence of caregiving models developed and tested in low and middle-income countries, we chose this model to maximize comparisons to the wide evidence from the U.S. and other higher income country contexts where this model has been employed. (Pearlin, 1999; Pearlin et al., 1990) This analysis focused on perceptions of alternative supports and services to offset this caregiving stress process.

This paper uses evidence from a mixed-methods study of caregivers of older adults in a city in northwest India (Jodhpur, Rajasthan) to address these gaps about long-term care needs and preferences. This paper has two aims:

1. Explore family caregivers’ perceptions about formal care services, and
2. Identify which kinds of LTSS caregivers report needing and are likely to use.

We address the first aim with qualitative data from in-depth interviews with caregivers (n=30) and the second aim with quantitative data from a survey of caregivers (n=100). Specifically, for the second aim, we describe stated preferences for long-term care services and test the hypothesis that family caregivers state a lower preference for formal care options than informal/in-home care options.

## Methods

This sequential mixed-methods study of caregivers for older adults was conducted in urban Jodhpur, the second largest city in its state (Rajasthan) and home to approximately 2.4 million residents, in two sequential waves – qualitative and then quantitative. Participants for both the qualitative and quantitative study were recruited from the geriatrics clinic at Mathura Das Mathur Hospital, a large public hospital affiliated with the largest medical college in Jodhpur. The geriatrics clinic was the second regional center for geriatrics excellence established as part of India’s National Program for Health Care of the Elderly.(Verma & Khanna, 2013) In line with the NPHCE goals, the clinic primarily serves as an outpatient clinic unit with dedicated clinicians, a pharmacy, and a laboratory to improve older adults’ quality of care at public hospitals. Although the Jodhpur clinic is a referral point for primary health centers from around the region, this study focused only on families within the limits of Jodhpur city.

The study population was adults (aged 18+) caring for a family member who was able to provide informed consent. Study staff introduced the study goals and procedures to potential participants when an older adult presented at the clinic. Those interested and eligible provided the telephone number of the primary caregiver for the older adult who came to the clinic. Study staff then called the primary caregiver, screened them for eligibility, and scheduled appointments with eligible participants upon commencement of data collection. The study was approved by the University of Minnesota in Minneapolis and Tata Institute of Social Sciences in Mumbai Institutional Review Boards, respectively.

### Qualitative Study Design

The qualitative wave conducted one-on-one, in-person, semi-structured interviews with 30 family caregivers stratified by care recipient condition. The semi-structured interview topics were informed by Pearlin’s stress and coping model, commonly invoked by caregiving studies in Western settings. (Pearlin, 1999; Pearlin et al., 1990) The data analysis was a mix of inductive and deductive coding.

#### Sampling Design and Study Recruitment

The study used a stratified, purposive sampling approach for caregivers of older adults. The sample was stratified *a priori* by intensity of care demands: (1) severe physical impairment or at least mild cognitive impairment; (2) mild-to-moderate physical impairment; (3) little-to-no physical impairment. The sample was stratified for two reasons. First, there is often a baseline of care or help that most older adults in India receive as a function of multigenerational living arrangements; older adults in homes with younger people would be unlikely to cook. Including the little-to-no care group was intended to represent this baseline care that older Indian adults receive and would serve as a comparison to the specific kinds of care for limitations and health conditions. Second, we included two categories of caregivers providing care to older adults with severe to mild impairments to capture the unique challenges posed as care needs progressively increase. The study was introduced to clinic patients in person while they were at the clinic between September and December 2014.

#### Data Collection

Study interviews were completed in the caregiver’s home after reviewing study procedures, and the caregiver provided informed consent to participate in the study. The semi-structured interviews covered several domains of caregiving responsibilities, including the temporal narrative of how care demands emerged, the caregiver’s current daily duties, and subjective processes of stress and coping. In line with Pearlin’s model, caregivers were asked throughout the semi-structured interview to offer insights into the care-recipient characteristics, and their experience of the objective instrumental and financial caregiving demands, other life demands, secondary role strain and benefit, caregiver physical and mental health, perceived social support, and family dynamics. In addition to suggested questions and anticipated relevant probes included in the interview guide, interviewers were encouraged to probe on relevant themes and topics to comprehensively explore the caregivers’ experience providing care.

The interviews lasted between 60-90 minutes and were conducted between December 2014 and February 2015. Two study research assistants were present at each interview: one who conducted the interview and the other who took notes to capture non-verbal communication and contextual insights that complemented the interview data. The interviews were conducted in the local language of the caregiver’s preference; the majority of interviews were conducted in Hindi and a minority were conducted in Marwadi, the local language in Jodhpur. Interviews were audio recorded, transcribed, and translated into English; a sub-sample of the translations was quality checked by an independent research assistant fluent in Hindi and Marwadi. Data analysis was conducted using the English transcripts.

#### Data Analysis

Data analysis was informed by thematic analysis approaches(Braun & Clarke, 2006). Inductive and deductive codes were applied based on the structure of the interview guide, our conceptual framework, and themes that emerged from the data. For this particular analysis, a hybrid approach of an *emic* process of coding new themes and an *etic* process of applying established Pearlin’s model(Leonard I. Pearlin et al., 1990) to further organize themes was used to maximize implications for future research and practice; emic themes that emerged from the data. Initial coding and subsequent organization of codes into hierarchical categories was conducted in Microsoft Excel. The codes were then grouped into hierarchical categories as a means of organizing the coded data, akin to parent/child nodes.(Corbin & Strauss, 2008) For example, codes such as “prayer” and “*dharma*” were clustered under a category of “religious explanations and coping behaviors. The coding process integrated insights and input from the investigator team, including the investigator who led the coding/categorization.

### Quantitative Study Design

The quantitative wave consisted of individual surveys with 100 family caregivers recruited from Mathura Das Mathur Hospital. The survey covered multiple domains, informed by both Pearlin’s model and evidence from the qualitative data. This analysis focuses specifically on a module of questions pertaining to preferences for additional supportive services to care for the older adult in their family for whom the respondent was currently providing care.

#### Study Sample, Recruitment, and Data Collection

The quantitative data comes from a clinic-based sample of patients who sought care at MDM Hospital’s Regional Centre for Geriatric Excellence between December 2014 and May 2015. When seeking care, patients and their caregivers were asked if they would be willing to be contacted about the study; if so, they provided their contact information to connect with the caregiver. Study recruitment began in April 2015, and potential caregivers were contacted and recruited to participate. In the case of multiple caregivers, the caregiver who was enrolled in the study was the patient’s *primary* caregiver as identified by the patient or household head.

Data collection consisted of an individual interview with the primary caregiver at the caregiver’s home. The interview involved several domains, including background demographic characteristics and socioeconomic status; perceptions of social norms for care for older adults; care recipient’s needs; network of all the patient’s caregivers and the kinds of care each provided; the care provided to other older adults in addition to the patient; caregiver physical well-being, physical health and mental health; caregiver social support; and preferences for and perceived unmet needs for additional supports to provide care to the patient. Interviews lasted about one hour and were conducted in Hindi or Marwadi. Respondents’ answers were recorded on paper surveys by the interviewer; data were entered separately into a database using a double-data entry approach. Missing data or enumeration errors identified during data entry were addressed by the interviewer/enumerator, including contacting the caregivers as needed. Data collection was completed by June 2015, and data entry, cleaning, and management were carried out through the end of 2015. Participant characteristics of the qualitative sample are presented in Table 1.

#### Service Needs Measure

Based on qualitative data, literature reviews, existing measures, and insights from local clinicians and key informants, we developed a series of questions to ascertain preferences for and likelihood of using supportive services to facilitate care for older adults. Specifically, the service needs and preferences module asked caregivers to report whether they needed a number of services (yes/no). If the caregiver reported that they needed the service, they were asked to respond on a four-point Likert scale (very unlikely to very likely): “how likely would you be to take advantage of this service if it was provided?” The specific items included respite care, stress management, learning more about care-related tasks, self-care, trained nursing/home health care staff, assistance with instrumental activities of daily living, medical social work/care management at the hospital, support group, financial help, instruments and devices (e.g., wheelchair). We also calculate the sum, or total number, of supportive services the caregiver reported needing.

#### Predictors of Supportive Service Needs and Preferences

For this analysis, we include additional factors captured in the survey that likely explain supportive service preferences and needs – caregiver characteristics and socioeconomic status, care recipient characteristics, caregiving burden and intensity, and caregiver health. Background characteristics included age (continuous, centered at mean), gender (binary, reference: male), and education (categorical: none, primary/middle school, secondary (reference), graduate/post-graduate). Care recipient characteristics included relationship to caregiver (categorical: spouse, parents, parents-in-law (reference), other family) and reasons for care (categorical: old age, physical health (reference), cognitive/mental health). Caregiving burden and intensity included the 12-item version of the Zarit Caregiver Burden Inventory(Bédard et al., 2001) (tertiles) and self-reported hours per week of care provided (continuous). Caregiver physical health was assessed with the physical components score of the 12-Item Short Form Health Survey (SF-12) (Ware, Kosinski, Turner-Bowker, & Gandek, 2005), where self-reported domains of physical health-related quality of life are scored on a scale of 0-100, with higher scores being higher quality of life.

#### Analysis

First, we described the prevalence of preference for individual supportive services as well as the number of supportive services reportedly wanted, as little is known about the population of older adult caregivers’ needs and preferences for long-term supportive services in India. To estimate the predictors of the extent of supportive services needed, we fit multivariable-adjusted Poisson regression models of the number of services required. Poisson models best fit the count nature of the dependent variable, the sum of the number of supportive services needed. To identify predictors of specific kinds of supportive services, we fit a series of logistic regression models of each kind of supportive service as dependent variables in its respective model. We also calculated and plotted predicted probabilities of reporting needing each of these kinds of supportive services; we estimated these probabilities in the full sample (i.e., not sex-stratified) because the frequency of needing some of the kinds of supportive services prevented fitting sex-stratified models in our relatively small sample.

## Results

### Sample characteristics

#### Qualitative sample characteristics

Caregivers were mostly women (28 of 30), average age of 49 years old. Most provided care for a mother-in-law (n=13) or spouse (n=8). Others were caring for their father-in-law (n=4), their mother (n=3), or a grandparent (n=2). Eighteen caregivers (60%) were providing physical care, six were providing normal aging care, and six were caring for elders with cognitive issues.

#### Quantitative sample characteristics

Characteristics of quantitative wave participants, stratified by gender, are presented in Table 2. The average participant was 50 years old with higher education, caring for a parent (men) or parent-in-law (women) with age-related limitations, low caregiver burden, high caregiver benefits, 60.8 hours per week, and with average HRQOL. As the HRQOL measure is normed to the American standard average on a 0-100 scale, all participants have lower than average HRQOL (<50).

The median number of LTSS caregivers needed was 2 (interquartile range: 0-5; Figure 1). Caregivers most commonly reported wanting self-care activities and care education; respite care, hospital services / medical social work services, financial help, and devices/instruments were the next most commonly reported LTSS, each reported as being needed by a similar number of caregivers (Figure 2). In bivariate analysis (t-test of number of LTSS needs; chi-square of frequency of specific LTSS needs), there were no significant differences in total number of *Study goal 1:*

**Table 1.**
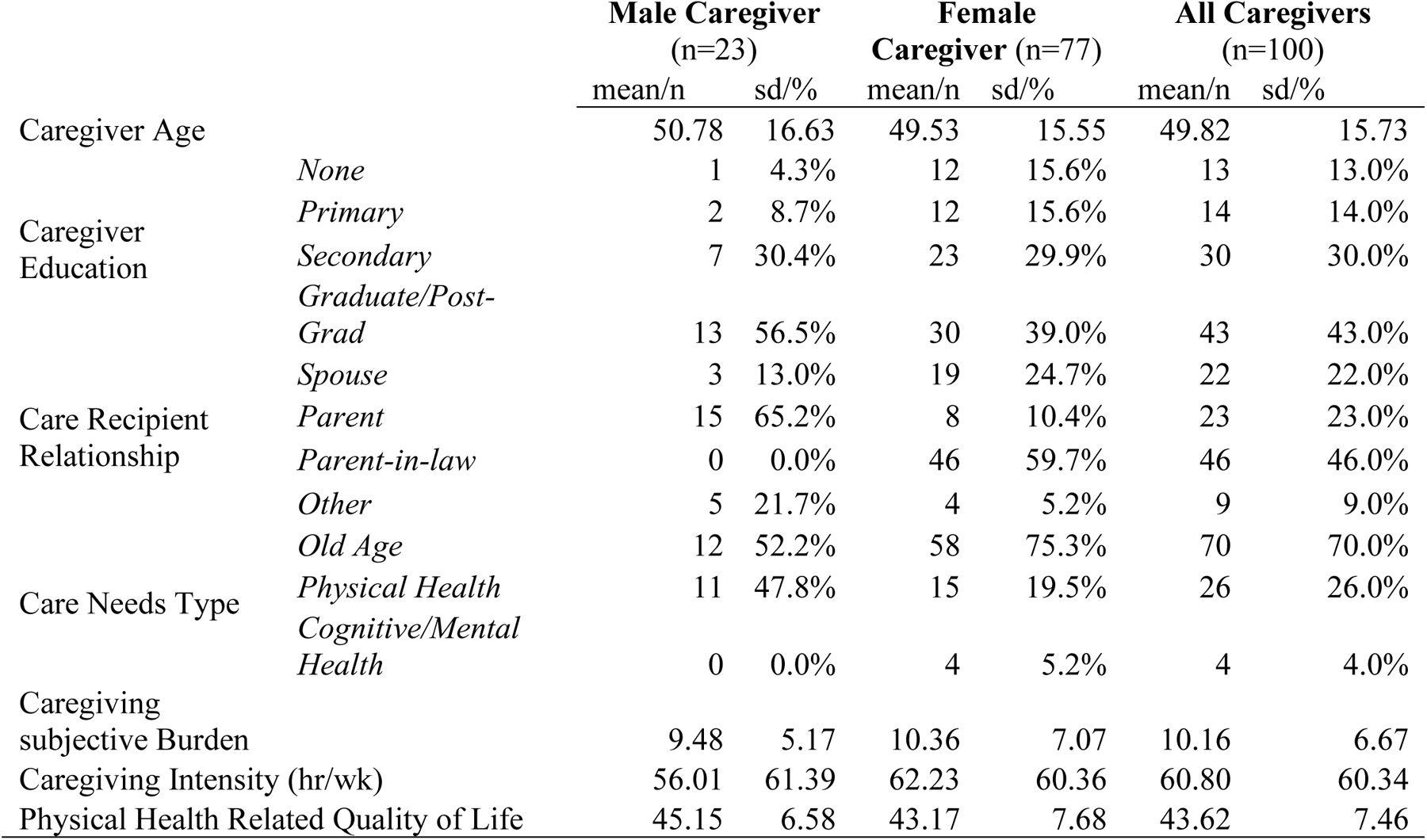
Characteristics of Quantitative Study Participants by Caregiver Sex.

**Figure 1.**
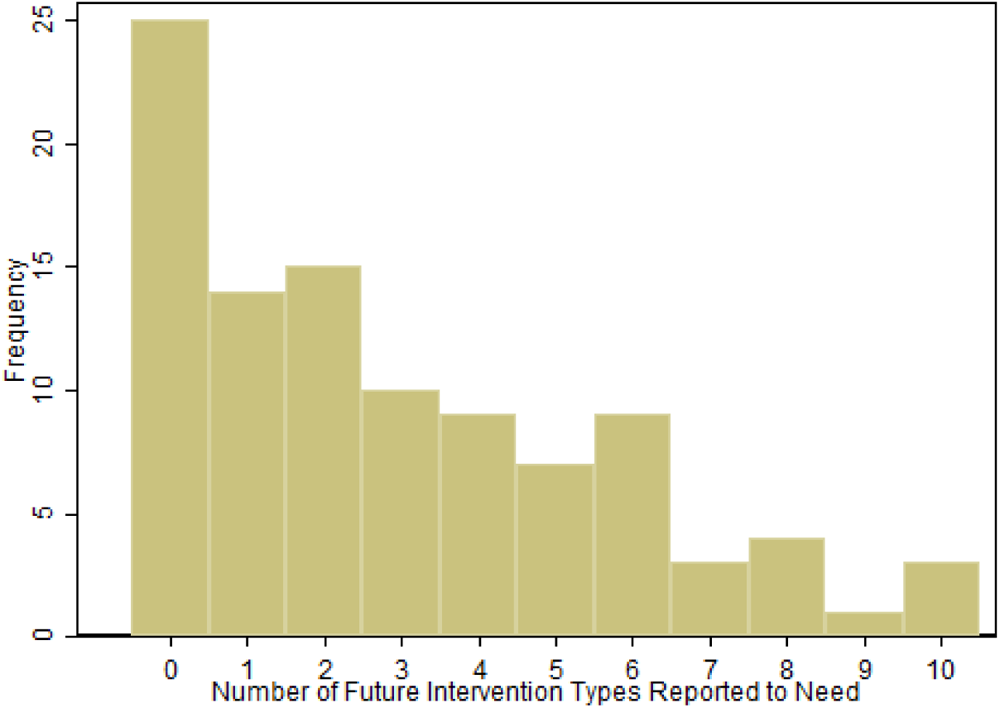
Distribution of Number of LTSS Needs Caregivers Reported

**Figure 2.**
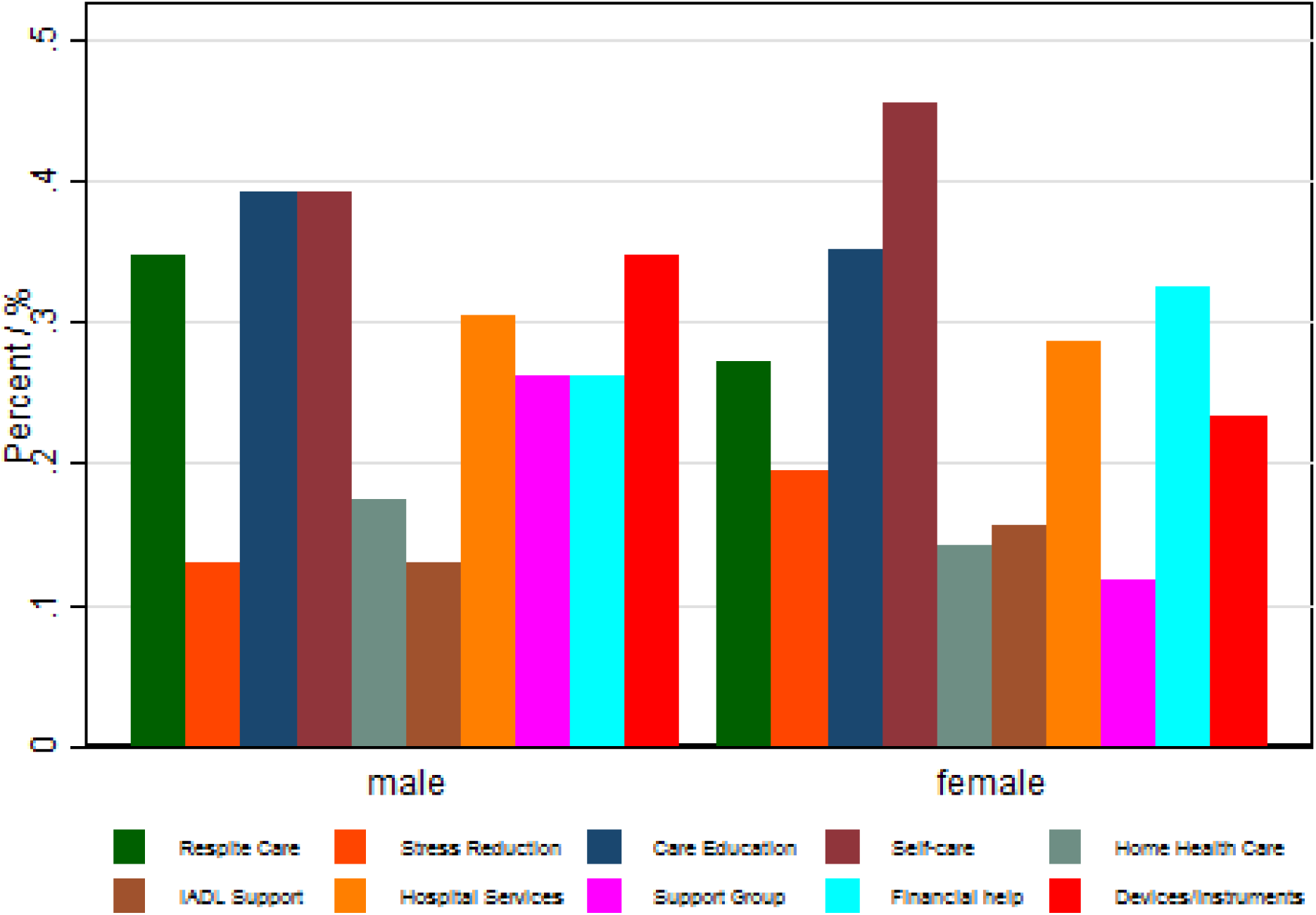
Prevalence of Individual LTSS Needs

The first goal of the study was to explore family caregivers’ perceptions about formal care services. The following are the main themes:

##### 1. Disinterest in formal care services

Caregivers typically stated themes of disinterest of formal care services, either of in-home or residential services. For the in-home case, one caregiver (“R”) articulated the theme that a paid helper providing care for money instead of by duty or affection was inherently lower quality in this exchange with the interviewer (“M”):

> R-If we would have had money, we would have hired a servant to make him get up and sit. Today the servant is asking for Rs.1500/-for each day. That means Rs.45,000/-so where from will we bring 48,000? We needed only for latrine etc. Like for bathing and washing him. Where from to bring this much money! So we cannot hire. Main is keeping the servant, what else? On contrary there is a risk in hiring a servant. I also cannot walk much so again and again I can’t go behind him/her, nowadays there is theft, so keeping him and then taking Acres of his food, etc, what is not there at home, so what will I know what they have carried away. What will I learn! So better than that we do ourselves. If I have to pay attention to him, better I pay attention to my father in law and take care of him.
>
> M-if you would not have to think about the money, would you have hired a servant then?
>
> R-No even then I would have not hired. How much the family member or blood relatives are able to take care, that much an outsider will not. The other person will do it for money but we do it with the affection towards him. So I would not hire. [ID13].

One caregiver succinctly summarized this sentiment: *“Any outsider cannot come and help you in this.” [ID24]*.

##### 2. Culture of care

There was also a strong theme of caregivers perceiving that they were the only people to provide care (“If I do not look after then who will do?” [ID03]). While this sentiment is imbued with themes of duty and reflects the caregiver’s motivation to provide care, this rhetorical question also reflects a perceived absence of alternatives. In other words, this common idea may simply reflect a lack of acknowledgement, understanding or availability of other sources of long-term support for this older adult. One caregiver articulated this idea about someone else from outside the family helping her provide this care, *“We do not think to take any one’s service. We do not keep any expectations. If they wish, they would do by themselves only. [We] cannot tell him to do it compulsorily.”* [ID20] Notably, for those in a joint family household (i.e., multi-generational household) the larger numbers of people nearby facilitate sharing care duties. “*In a joint family, it is easier to take care. Suppose you have to go somewhere out, whoever is living with you, will help you in taking care of the person. So you can go out relaxingly if you have to.” [ID04] Some caregivers noted unpaid help from neighbors as a source of informal support*.

For some hiring to provide formal care services would reflect that they were providing insufficient or inadequate quality care. *“According to me I have given 100% to paternal grandfather in law. If something is lacking then [it is my fault].” [ID09]*.

##### 3. Distrust in paid care

Domestic help (“servant”), is not uncommon in Indian families with financial means to hire help for cooking and cleaning, could also theoretically be tasked with personal care with activities of daily living for the care recipient. This caregiver also notes concerns about safety, theft and oversight of this servant. There is also a concern about paid caregivers not doing or doing inferior quality work, in part because they were doing the work for money. This sentiment about distrust of paid work was common: “We take them to the hospital and whatever service is there, we do. We are not leaving them on the trust of others that the other person will do. We do ourselves. We are not trusting others.” [ID15].

As such family caregivers perceive the benefit of providing care by themselves. As note by a respondent caring for her parents-“*The biggest benefit is trust, I have that satisfaction that my parents are with me. They also have that faith in me that I am there for them and whenever they will call me I shall be there for them.” [ID 27]*.

##### 4. Hiring to avail expert medical care

A small minority of families did use formal health care services, like home health care nursing staff, and those who did reported a theme of *expertise* by these providers. A number these caregivers mentioned physiotherapists as valuable formal medical care options that came into their home, though arguably distinct from long-term services and supports. In one case, a male caregiver was very intentional about wanting to learn from the nursing aid the more technical skills (wound care for bed sores) so that he could provide better support when, inevitably, there came a time when the nursing staff could not be there to do it themselves.

Another woman who was providing care for her spouse who was bedridden and severely physically limited had in-home nursing support and medicalized arrangements in the home (“The room is like an ICU:”). She notes a similar sense of expertise in the nursing staff, particularly in the context of triaging care and interfacing with doctors: “*The nursing staff here is so expert that they know what treatment in emergency is needed and they provide it. They talk to the doctor and ask as what is to be given.” She reports a similar theme of gratitude for such service, noting that her sons are able to pay: “Yes, all cannot have [nursing staff in home]. We are lucky that our children are so able. Everything depends on the money. Everyone is not getting all this so they have to do themselves.” [ID06]* A few (<5) caregivers were either themselves nurses or had a neighbor and family members who are nurses who could provide more medicalized care for free [ID08]. Other than servants or domestic helpers, only one caregiver noted paying for non-professional supportive services – she would pay a nominal amount to taxi drivers who waited nearby while not driving, to help transfer her mother-in-law in and out of a wheelchair to get her to the toilet. [ID16]

Some caregivers did note services and supports that would be useful; however, many caregivers had difficulty conceiving of what they might want or need and stated that they wish for the care recipient’s condition or difficulties to resolve. Many reference wheelchairs, canes, walkers and other mobility aids as being helpful. One caregiver mentioned that getting her mother in and out of the house to go to the hospital as particularly challenging: “When my children are not at home, I have to call neighbor…. I will make [mother] sit on a chair and [the neighbor and I] both together will carry her out or to take her to hospital if something happens.” [ID08] A number of caregivers mentioned the usefulness of machines to help with caregiving – massage machines, blood pressure or blood sugar monitors. Others noted transportation more broadly as an issue, especially taking a care recipient with mobility difficulties to the hospital. If families did not have a car or bicycle available, they had to find and hire an auto-rickshaw; others noted the cost of transportation to the hospital as challenging. Financial assistance for medications was nearly universally mentioned.

*Study goal 2:*

The second goal of the study was to identify which kinds of services caregiver’s report needing and likely to use. Female caregivers reported requiring significantly fewer total LTSS services than men (beta:-0.415, p<0.01, Table 2). This is reflected in the differences in in the intercepts in the gender stratified models, where female caregivers have a lower intercept of LTSS services needed (1.378) than men (2.942). (Caregiver age was inversely associated with needing LTSS; each year of age was associated with fewer LTSS needs (beta: -0.01 (p<0.001), which was a small but statistically significant association. Caregivers providing health related care reported higher number of LTSS needs than those providing care for reasons of ‘normal aging (beta:0.854, p<0.001). Care hours per week were positively associated with LTSS services needed (beta: 0.00224, p<0.05); in other words, caregivers providing higher amounts of hours of care per week reported having more LTSS needs than caregivers proving fewer care hours per week. Caregiver health was inversely associated with LTSS; those with better HRQOL reported fewer LTSS need (beta: -0.02, p<0.01). Perceived caregiving burden was inversely associated with LTSS needs for women (p<0.05) and positively, though not statistically significantly, associated with LTSS need for men.

**Table 2.**
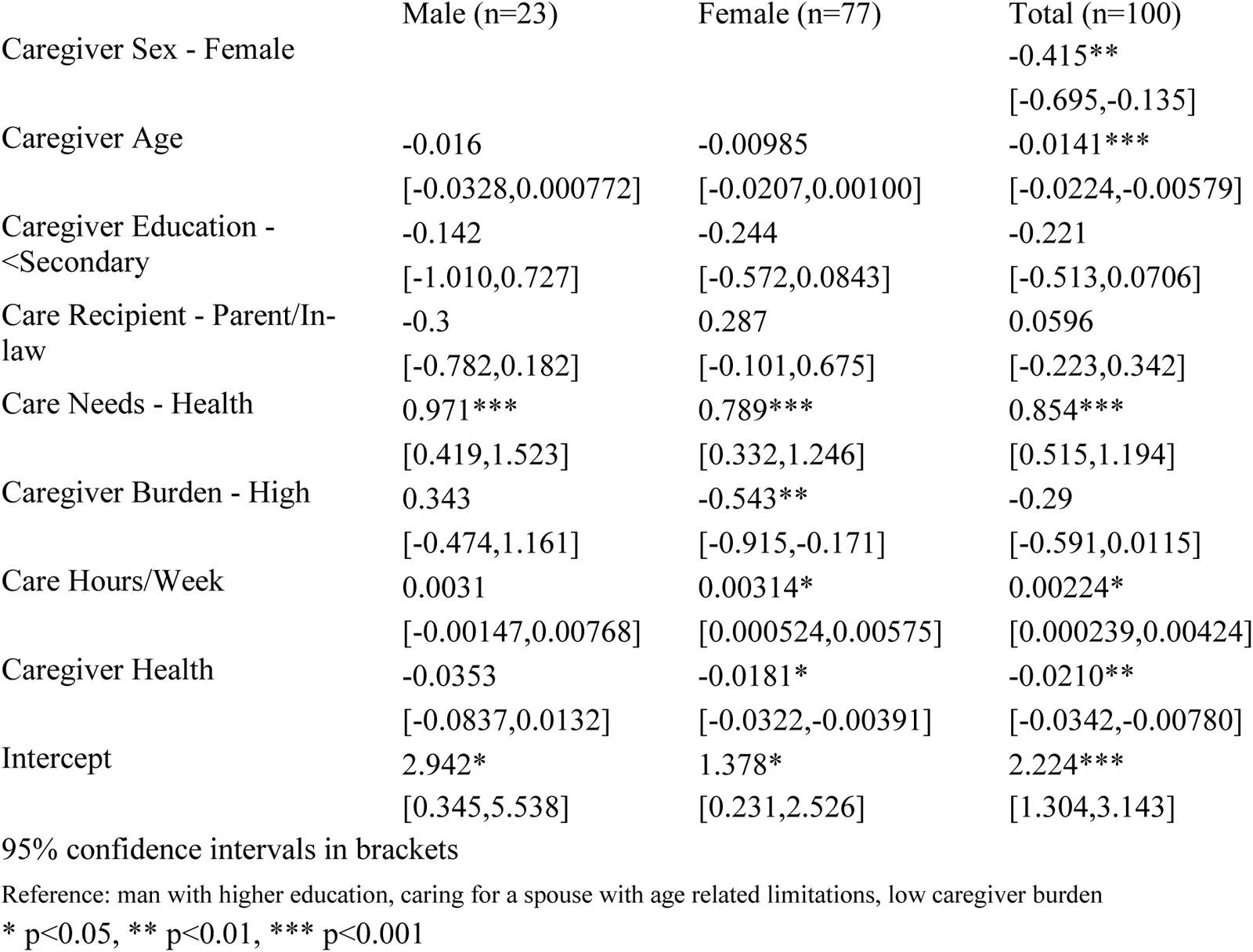
Association of Caregiver and Care Recipient Factors and Number of Identified Long Term Supports and Services.

In general, providing health related caregiving was associated with higher odds of needing each of the individual LTSS compared to those providing care for “normal” aging older adults (Table 3). Older age was generally associated with a lower odd of needing each of the caregiving services, notably care education, support groups, and financial support, respectively (p<0.05). Female caregivers had a lower odd of needing LTSS’s such as financial support (OR: 0.26, p<0.05) than male caregivers. Higher caregiver burden was associated with lower odds of needing support groups and stress reduction (OR: 0.24, p<0.05 for each) than caregivers with low burden. Caregiver health was associated with lower odds of LTSS needs, especially care education (OR:0.92, p<0.05) and home health care (OR:0.88, p<0.05). Although the coefficients were not statistically significant, those with lower education had lower odds of needing supportive services, except for financial support and devices where they had a higher odd, compared to those with higher education.

**Table 3.** Odds of Needing Long Term Supports and Services.

|  | Respite | Stress<br>Reducti<br>on | Care<br>Education | Self<br>Care | Home<br>Health<br>Care | IADL<br>Health | Hospital<br>Support | Support<br>Group | Financial Support |
| --- | --- | --- | --- | --- | --- | --- | --- | --- | --- |
|  | OR<br>[95%<br>CI] | OR<br>[95%<br>CI] | OR [95%<br>CI] | OR<br>[95%<br>CI] | OR<br>[95% CI] | OR [95%<br>CI] | OR [95%<br>CI] | OR [95%<br>CI] | OR [95% CI] |
| Caregiver<br>Sex –<br>Female | 0.462<br>[0.149,1<br>.435] | 0.807<br>[0.263,2<br>.475] | 0.783<br>[0.251,2.44<br>1] | 0.832<br>[0.195,3<br>.548] | 0.813<br>[0.167,3.<br>948] | 0.601<br>[0.179,2.0<br>16] | 0.295<br>[0.0725,1.<br>203] | 0.714<br>[0.200,2.5<br>57] | 0.267*<br>[0.0754,0.944] |
| Caregiver<br>Age | 1.001<br>[0.969,1<br>.034] | 0.971<br>[0.940,1<br>.003] | 0.960*<br>[0.928,0.99<br>3] | 1.024<br>[0.978,1<br>.072] | 0.985<br>[0.942,1.<br>031] | 0.973<br>[0.940,1.0<br>07] | 0.968<br>[0.929,1.0<br>08] | 0.957*<br>[0.923,0.9<br>93] | 0.964*<br>[0.931,0.998] |
| Caregiver<br>Education -<br><Secondary | 0.765<br>[0.252,2<br>.324] | 0.387<br>[0.123,1<br>.217] | 0.565<br>[0.188,1.69<br>5] | 0.223<br>[0.0364,<br>1.367] | 0.636<br>[0.135,2.<br>995] | 0.642<br>[0.201,2.0<br>47] | 0.353<br>[0.0618,2.<br>015] | 2.265<br>[0.722,7.1<br>08] | 1.529<br>[0.489,4.780] |
| Care<br>Recipient -<br>Parent/In-law | 0.976<br>[0.317,3<br>.006] | 1.273<br>[0.420,3<br>.854] | 1.294<br>[0.419,3.99<br>7] | 0.601<br>[0.134,2<br>.692] | 2.305<br>[0.466,1<br>1.40] | 0.698<br>[0.220,2.2<br>14] | 0.263<br>[0.0632,1.<br>096] | 0.454<br>[0.133,1.5<br>48] | 0.462<br>[0.144,1.476] |
| Care Needs<br>– Health | 3.513<br>[0.989,1<br>2.48] | 2.524<br>[0.802,7<br>.947] | 5.540**<br>[1.661,18.4<br>7] | 3.709<br>[0.666,2<br>0.67] | 2.196<br>[0.371,1<br>3.00] | 5.620*<br>[1.315,24.<br>03] | 6.379<br>[0.916,44.<br>40] | 13.70**<br>[2.264,82.<br>88] | 6.436*<br>[1.360,30.47] |
| Caregiver<br>Burden –<br>High | 1.141<br>[0.379,3<br>.439] | 0.238*<br>[0.0664,<br>0.857] | 0.606<br>[0.197,1.86<br>4] | 0.509<br>[0.105,2<br>.456] | 0.149<br>[0.0150,<br>1.483] | 0.268<br>[0.0681,1.<br>056] | 1.268<br>[0.264,6.0<br>97] | 0.238*<br>[0.0588,0.<br>959] | 0.814<br>[0.231,2.868] |
| Care<br>Hours/Week | 1<br>[0.992,1<br>.008] | 0.999<br>[0.990,1<br>.007] | 1.001<br>[0.993,1.00<br>9] | 1.006<br>[0.996,1<br>.016] | 0.982<br>[0.964,1.<br>001] | 1.001<br>[0.992,1.0<br>10] | 1.003<br>[0.993,1.0<br>14] | 1.002<br>[0.993,1.0<br>12] | 1.002<br>[0.994,1.011] |
| Caregiver<br>Health | 0.958<br>[0.898,1<br>.021] | 0.981<br>[0.919,1<br>.047] | 0.922*<br>[0.850,0.99<br>9] | 0.965<br>[0.900,1<br>.035] | 0.878*<br>[0.780,0.<br>989] | 0.923<br>[0.849,1.0<br>02] | 1.082<br>[0.960,1.2<br>20] | 0.971<br>[0.910,1.0<br>37] | 0.951<br>[0.890,1.016] |

Reference: man with higher education, caring for a spouse with age related limitations, low caregiver burden
\* $p<0.05$ , \*\* $p<0.01$ , \*\*\* $p<0.001$

## Discussion

This mixed-methods study of family caregivers for older adults in Jodhpur, Rajasthan found strong evidence of a cultural relevance for family members providing long-term supports and services for older adults but also unmet needs for additional services to assist in this care. Both the qualitative and quantitative data showed in home health care and domestic labor (IADL care in the quantitative survey) were the least frequently reported LTSS needs. Indeed, self-care and care education, which would facilitate caregivers to carry out their caregiving roles in home, were the most commonly reported LTSS needs. In particular, we found evidence that women reported fewer needs for supportive services and that providing health related care was associated with significantly higher odds of reporting LTSS than “normal aging” care.

### Strengths and Limitations

While this study offers unique insights into the LTSS preferences and needs of caregivers for older adults in India, our findings should be considered in light of the study limitations. Our stratified sampling for the qualitative sample was unbalanced more than was planned; there were also unanticipated blending between caregiving groups (i.e., those with physical and cognitive issues). Although we had sufficient sample to reach saturation within the normal aging and cognitive caregivers, we do not make formal comparisons across these groups in this analysis for brevity. We did not stratify the qualitative sample by caregiver sex; for this analysis, this may have resulted in us missing some of the LTSS that men would have wanted in the quantitative survey because we did not capture their needs first qualitatively. In the quantitative sample, we note that the convenience sample and cross-sectional design limit generalizability and our ability to identify temporality, respectively. The quantitative sample’s size, while sufficient for crucial statistical assumptions like the central limit theorem, limited extensive modeling of LTSS needs as outcomes. Lastly, we note that our primary survey questions for this work were from a newly designed measure for this population and thus were not previously validated.

These limitations notwithstanding, our study has a number of strengths. It is one of the first mixed methods studies of caregivers’ LTSS needs and preferences in India. The richness of the qualitative data helped inform a tailored survey measure of relevant LTSS for this population. Moreover, the combination of results from qualitative and quantitative waves strengthens the validity of our findings. The classification of “normal aging” related care and health related care facilitated comparing the needs of those providing health-specific caregiving to the typical expectations of older adults support common in Jodhpur.

### Comparisons with Prior Literature

These results suggested that caregivers prioritized LTSS, like self-care and care education, that complement or facilitate their caregiving role and expressed disinterest in formal in-home LTSS like nursing staff or domestic help that would substitute their caregiving roles. Other studies of caregiving in India have found most care comes from families.(Berkman, Sekher, Capistrant, & Zheng, 2012; Chakraborty & Bansod, 2014; Gupta, 2009; Gupta, Rowe, & Pillai, 2009; V. Patel & Prince, 2001) One explanation for this result is the cultural meaning attributed to providing caregiving; caregivers actively want to provide this care as an expression of love, duty, and honor to their elder.(S. Lamb, 2005; S. E. Lamb, 2009) The cultural meaning may be socially enforced – that formal support reflects the family abdicating their responsibility and may result in negative standing within extended family or neighbors --(S. Lamb, 2013) and even through cultural nostalgia in media.(Dey, 2016) This process may also stem from misunderstandings and under-diagnosis of conditions in later life, like dementia, as medical conditions that are beyond the scope of “normal aging.”(Patel & Prince, 2001) It is thus possible that lower support for formal LTSS results from a lower perception of the care needs as medical needs that requires specialized care. It is worth noting that caregivers providing health-related care were more likely to report needing these services; caregivers who perceive their care as due to a health condition rather than a function of old age may have lower stigma about available formal services.

Female caregivers reported lower number of LTSS needs and having lower odds of reporting specific LTSS needs than men. This quantitative finding might connect with the largely female qualitative sample where some caregivers were not necessarily aware or communicative of their needs. It is possible that women have lower autonomy, bargaining power or resources get support the need (Bloom, Wypij, & Das Gupta, 2001; Malhotra, Malhotra, Ostbye, & Subramanian, 2014; Pezzin & Schone, 1999) – this theme is reflected in the examples of caregivers with formal supports whose sons are able to pay – though this idea is speculative in our data.

The patterns of the associations’ direction were consistent with a caregiver healthy-worker selection process; that caregivers are those who are healthy enough to provide care.(Bertrand et al., 2012) Higher HRQOL was associated with lower odds of reporting needing care education and home health care suggests that those caregivers are healthy and able to sustain this extent of caregiving without perceiving need for supplemental LTSS.

### Implications for Policy, Practice and Future Research

These findings suggest that additional research is needed on the preferences for LTSS in India to inform relevant and appropriate health care and social policy investments to support family caregivers and care recipients. Others have called for an increase in research on LTSS in lower and middle-income country settings generally(Lloyd-Sherlock, 2014, 2015) and in India in particular.(Brijnath, 2012; Evans, Kiran, & Bhattacharyya, 2011) Specifically, future observational research from large, population-based surveys about needs and preferences for formal and informal caregiving, experimental studies of effective models of effective interventions to address these LTSS needs and preferences are also needed; and translational research to scale and roll out effective interventions in community settings(Dias et al., 2008) are all needed.

Civil society and health sector investments should prioritize services that support and facilitate caregivers to remain in their caregiving roles at home. In particular, care education and training in self-care activities appear most relevant for caregivers; evidence-based models should be developed and implemented in clinical and community settings. Indeed, it might be relevant to identify the ideal way to frame or brand such services to avoid stigma of formal LTSS. Many caregivers reported needing devices, especially wheelchairs; programs that could provide such devises at a low or no cost would be highly valued. Clinical practitioners’ should prioritize clear communication about prognosis after discharge for geriatric populations to help families identify the extent to which they need support.

### Conclusions

In sum, this mixed-methods study of Indian family caregivers’ perceptions and preferences of LTSS needs showed evidence that caregivers prefer LTSS that complement, not substitute, their roles as caregivers. Caregivers expressed themes of distrust of formal, paid help, that formal help was of lower quality; caregivers expressed themes of duty and affection. Those who did have formal help had someone in the family paying for it, or some hybrid where they had a nurse in the family who could provide highly skilled informal help. Self-care and care education were most commonly reported LTSS needs; home health care and domestic help were the least commonly reported needs. Caregivers for physical health conditions had significantly higher needs than caregivers or “normal” aging, and women reported fewer LTSS needs than men. Additional research and evidence-based practice work to provide caregivers with the kinds of LTSS they need. India’s demographic and epidemiologic transitions require attention on this issue to support families caring for older adults with more care needs in the coming decades.

## Data Availability

All data produced in the present study are available upon reasonable request to the authors

## Acknowledgement

The study was funded by Global Spotlight International Research Seed Grant, University of Minnesota Twin Cities. We are thankful to Bea Capistrant, VP of Research, JP Morgan Chase who served as the Co-PI on this project.

## REFERENCES

Apt, N. A., & Gricco, M. (1994). Urbanization, caring for elderly people and the changing African family: The challenge to social policy. International Social Security Review, 47(3-4), 111–122.

Balarajan, Y., Selvaraj, S., & Subramanian, S. V. (2011). Health care and equity in India. [Research Support, N.I.H., Extramural Research Support, Non-U.S. Gov’t]. Lancet, 377(9764), 505–515. doi: 10.1016/S0140-6736(10)61894-6

Bédard, M., Molloy, D. W., Squire, L., Dubois, S., Lever, J. A., & O’Donnell, M. (2001). The Zarit Burden interview a new short version and screening version. The Gerontologist, 41(5), 652–657.

Berkman, L. F., Sekher, T., Capistrant, B. D., & Zheng, Y. (2012). Social Networks, Family, and Care Giving Among Older Adults in India. In J. P. Smith & M. Majmundar (Eds.), Aging in Asia: Findings from New and Emerging Data Initiatives (pp. 261–278). Washington, DC: The National Academies Press.

Bertrand, R. M., Saczynski, J. S., Mezzacappa, C., Hulse, M., Ensrud, K., & Fredman, L. (2012). Caregiving and cognitive function in older women: evidence for the healthy caregiver hypothesis. [Research Support, N.I.H., Extramural]. J Aging Health, 24(1), 48–66. doi: 10.1177/0898264311421367

Bloom, S. S., Wypij, D., & Das Gupta, M. (2001). Dimensions of women’s autonomy and the influence on maternal health care utilization in a north Indian city. . Demography, 38(1), 67–78.

Braun, V., & Clarke, V. (2006). Using thematic analysis in psychology. Qualitative Research in Psychology, 3(2), 77–101. doi: 10.1191/1478088706qp063oa

Brijnath, B. (2012). Why does institutionalised care not appeal to Indian families? Legislative and social answers from urban India. Ageing and Society, 32(04), 697–717.

Chakraborty, S., & Bansod, D. (2014). Scion’s Care Meliorates Elderly Health: A Study of Differential in the Care and Support and its Impact on Wellbeing of Elderly in India. Journal of Asia Pacific Studies, 3(3), 300–337.

Corbin, J. M., & Strauss, A. L. (2008). Basics of qualitative research : techniques and procedures for developing grounded theory (3rd ed. ed.). Los Angeles, CA: Los Angeles, CA. : Sage Publications, Inc.

Dey, D. (2016). The Nostalgia of Values: Popular Depictions of Care Crisis towards Ageing Parents in India. Journal of Human Values, 22(1), 26–38. doi: 10.1177/0971685815608060

Dias, A., Dewey, M. E., D’Souza, J., Dhume, R., Motghare, D. D., Shaji, K. S., . . . Patel, V. (2008). The effectiveness of a home care program for supporting caregivers of persons with dementia in developing countries: a randomised controlled trial from Goa, India. [10.1371/journal.pone.0002333]. PLoS ONE, 3, e2333–e2333.

Duran, A., Kutzin, J., & Menabde, N. (2014). Universal coverage challenges require health system approaches; the case of India. Health policy, 114(2), 269–277.

Evans, J. M., Kiran, P. R., & Bhattacharyya, O. K. (2011). Activating the knowledge-to-action cycle for geriatric care in India. [journal article]. Health Research Policy and Systems, 9(1), 1–10. doi: 10.1186/1478-4505-9-42

Gupta, R. (2009). Systems perspective: understanding care giving of the elderly in India. Health Care for Women International, 30(12), 1040–1054.

Gupta, R., Rowe, N., & Pillai, V. K. (2009). Perceived Caregiver Burden in India. [10.1177/0886109908326998]. Affilia, 24(1), 69–79.

Kalavar, J. M., Jamuna, D., & Ejaz, F. K. (2013). Elder Abuse in India: Extrapolating From the Experiences of Seniors in India’s “Pay And Stay” Homes. Journal of Elder Abuse & Neglect, 25(1), 3–18. doi: 10.1080/08946566.2012.661686

Kumar, A. K. S., Chen, L. C., Choudhury, M., Ganju, S., Mahajan, V., Sinha, A., & Sen, A. (2011). Financing health care for all: challenges and opportunities. The Lancet, 377(9766), 668–679.

Lamb, S. (2005). Cultural and moral values surrounding care and (in) dependence in late life: reflections from India in an era of global modernity. Care Management Journals, 6(2), 80–89.

Lamb, S. (2013). In/dependence, intergenerational uncertainty, and the ambivalent state: Perceptions of old age security in India. South Asia: Journal of South Asian Studies, 36(1), 65–78.

Lamb, S. E. (2009). Aging and the Indian diaspora: *Cosmopolitan families in India and abroad*: Indiana University Press.

Lloyd-Sherlock, P. (2000). Old Age and Poverty in Developing Countries: New Policy Challenges. World Development, 28(12), 2157–2168.

Lloyd-Sherlock, P. (2002). Formal Social Protection for Older People in Developing Countries: Three Different Approaches. Journal of social policy, 31(04), 695–695.

Lloyd-Sherlock, P. (2014). Beyond neglect: Long-term care research in low and middle income countries. International Journal of Gerontology, 8(2), 66–69.

Lloyd-Sherlock, P. (2015). Barriers to linking research and policy: the case of long-term care in low and middle income countries. Population Horizons, 12(2), 62–67.

Malhotra, C., Malhotra, R., Ostbye, T., & Subramanian, S. V. (2014). Maternal autonomy and child health care utilization in India: results from the National Family Health Survey. Asia Pac J Public Health, 26(4), 401–413. doi: 10.1177/1010539511420418

Patel, V., Parikh, R., Nandraj, S., Balasubramaniam, P., Narayan, K., Paul, V. K., . . . Reddy, K. S. (2015). Assuring health coverage for all in India. The Lancet, 386(10011), 2422–2435.

Patel, V., & Prince, M. (2001). Ageing and mental health in a developing country: who cares? Qualitative studies from Goa, India. Psychological Medicine, 31, 29–38.

Pati, S., Agrawal, S., Swain, S., Lee, J. T., Vellakkal, S., Hussain, M. A., & Millett, C. (2014). Non communicable disease multimorbidity and associated health care utilization and expenditures in India: cross-sectional study.. BMC Health Serv Res, 14, 451. doi: 10.1186/1472-6963-14-451

Pearlin, L. I. (1999). The stress process revisited. In J. C. Phelan & C. S. Aneshensel (Eds.), Handbook of the sociology of mental health (pp. 395–415). New York, NY: Klower Academic/Plenum Publishers.

Pearlin, L. I., Mullan, J. T., Semple, S. J., & Skaff, M. M. (1990). Caregiving and the Stress Process: An Overview of Concepts and Their Measures. The Gerontologist, 30(5), 583–594.

Pezzin, L. E., & Schone, B. S. (1999). Intergenerational household formation, female labor supply and informal caregiving: A bargaining approach. Journal of Human Resources, 475–503.

Powell-Jackson, T., Acharya, A., & Mills, A. (2013). An assessment of the quality of primary health care in India. Economic & Political Weekly, 48(19), 53–61.

Prinja, S., Aggarwal, A. K., Kumar, R., & Kanavos, P. (2012). User charges in health care: evidence of effect on service utilization & equity from north India. [Research Support, Non-U.S. Gov’t]. Indian J Med Res, 136(5), 868–876.

Prinja, S., Bahuguna, P., Pinto, A. D., Sharma, A., Bharaj, G., Kumar, V., . . . Kumar, R. (2012). The cost of universal health care in India: a model based estimate. PLoS ONE, 7(1), e30362.

Reddy, K. S., Patel, V., Jha, P., Paul, V. K., Kumar, A. K., & Dandona, L. (2011). Towards achievement of universal health care in India by 2020: a call to action.. Lancet, 377(9767), 760–768. doi: 10.1016/S0140-6736(10)61960-5

Shrestha, L. B. (2000). Population aging in developing countries. Health affairs, 19(3), 204–212.

van Doorslaer, E., O’Donnell, O., Rannan-Eliya, R. P., Somanathan, A., Adhikari, S. R., Garg, C. C., . . . Zhao, Y. (2006). Effect of payments for health care on poverty estimates in 11 countries in Asia: an analysis of household survey data. [Research Support, Non-U.S. Gov’t]. Lancet, 368(9544), 1357–1364. doi: 10.1016/S0140-6736(06)69560-3

Verma, R., & Khanna, P. (2013). National Program of Health-Care for the Elderly in India: A Hope for Healthy Ageing. Int J Prev Med, 4(10), 1103–1107.

Wachter, K. W. (1997). Kinship resources for the elderly. Philosophical Transactions of the Royal Society of London. Series B: Biological Sciences, 352(1363), 1811–1817.

Ware, J. E., Kosinski, M., Turner-Bowker, D. M., & Gandek, B. (2005). How to score version 2 of the SF-12 health survey (with a supplement documenting version 1): Quality Metric Incorporated.

. World Population Prospects: The 2015 Revision. (2015). In U. P. Department (Ed.), (Vol. 2015): United Nations, Department of Economic and Social Affairs, Population Division.

